# Genetic risk, parental history, and suicide attempts in a diverse sample of US adolescents

**DOI:** 10.1101/2022.06.11.22276280

**Authors:** Ran Barzilay, Elina Visoki, Laura M Schultz, Varun Warrier, Nikolaos P Daskalakis, Laura Almasy

**Affiliations:** Department of Child and Adolescent Psychiatry and Behavioral Science, Children’s Hospital of Philadelphia (CHOP), Philadelphia, US; Lifespan Brain Institute of CHOP and Penn Medicine, Philadelphia, US; Department of Psychiatry, Perelman School of Medicine, University of Pennsylvania, Philadelphia, US; Department of Biomedical and Health Informatics, Children’s Hospital of Philadelphia (CHOP), Philadelphia, PA, USA; Department of Psychiatry, University of Cambridge, Cambridge, United Kingdom; Department of Psychiatry, McLean Hospital, Harvard Medical School, Belmont, MA, USA; Stanley Center for Psychiatric Research, Broad Institute of MIT and Harvard, Cambridge, MA, USA; Department of Genetics, Perelman School of Medicine, University of Pennsylvania, Philadelphia, PA, USA

**Keywords:** Suicide attempt, genetics, polygenic risk prediction, family history, adolescents

## Abstract

**Background:** Adolescent suicide is a major health problem in the US marked by a recent increase in Black/African American youth suicide trends. While genetic factors partly account for familial transmission of suicidal behavior, it is not clear whether polygenic risk scores of suicide attempt have clinical utility in youth suicide risk classification.

**Objectives:** To evaluate the contribution of a polygenic risk score for suicide attempt (PRS-SA) in explaining variance in suicide attempt by early adolescence.

**Methods:** We studied N=5,214 non-related Black and White youth from the Adolescent Brain Cognitive Development (ABCD) Study (ages 8.9-13.8 years) who were evaluated between 2016 and 2021. Regression models tested associations between PRS-SA and parental history of suicide attempt/death with youth-reported suicide attempt. Covariates included age, sex, and race.

**Results:** Over three waves of assessments, 182 youth (3.5%) reported a past suicide attempt, with Black youth reporting significantly more suicide attempts than their White counterparts (6.1% vs 2.8%, P<.001). PRS-SA was associated with suicide attempt (odds ratio [OR]=1.3, 95% confidence interval [CI] 1.1-1.5, P=.001). Inclusion of PRS-SA explained 2.7% of the variance in suicide attempts, significantly more than the base model including only age, sex and race, which explained 1.9% of the variance (P=.001). Parental history of suicide attempt/death was also associated with youth suicide attempt (OR=2.9, 95%CI 1.9-4.4, P<.001). Addition of PRS-SA to the model that included parental history significantly increased the variance explained from 3.3% to 4% (P=.002).

**Conclusions:** Findings suggest that PRS-SA may be useful for suicide risk classification in diverse youth.

**Contribution to the Field Statement:** Adolescent suicidal behavior is a major health problem, with suicide being the 2^nd^ leading cause of death in youth. Research that improves our understanding regarding drivers of suicide risk in youth can inform youth suicide prevention strategies. Family history of suicide is an established risk factor for youth suicidal behavior. Current methods in psychiatric genetics allow calculation of polygenic risk scores that represent genetic liability to specific conditions. It is not clear whether polygenic risk score of suicide attempt can assist in risk classification, beyond family history. In this work, we show that in a sample of 5,214 youth ages 9-13, of which 3.5% reported past suicide attempt, polygenic score of suicide attempt was associated with youth suicide attempt. This association additively explained variance over and above parental history of suicide attempt/death. Findings make a case for the potential utility of incorporating polygenic risk scores as part of suicide attempt risk classification in youth, and suggest that polygenic scores may reveal genetic liability that is not captured by family history of suicide.

## Introduction

Suicide is the second leading cause of death in US adolescents (1). The rising rates of suicide among Black or African American youth is especially concerning (2). Suicide attempt is a complex behavior driven by genetic and environmental factors (3). Clinicians often use parental history of suicide attempt/death to estimate suicide risk (4). The potential of using polygenic scores of psychiatric phenotypes to assess genetic suicide risk is uncertain (5). It is not known whether polygenic score of suicide attempts (PRS-SA) can contribute to suicide risk classification, and whether PRS-SA adds useful information beyond the commonly used risk assessment based on parental history.

The Adolescent Brain Cognitive Development (ABCD) Study follows diverse genotyped US youth from ages 9-10 into adolescence (6). The study collects data on parental history of psychiatric conditions (7), including suicide attempt/death. Participants are evaluated annually for history of suicide attempts, and endorsement of suicide attempts in Black participants is significantly higher (8). Here we aimed to evaluate the contribution of PRS-SA in explaining variance in self-reported suicide attempt by early adolescence, and to determine the additive effect of this score over and above parental history of suicide attempt/death.

## Methods

### Participants

We included N=5,214 non-related ABCD Study participants of African and European ancestry (based on genetic principal components (9)) that had data on parental history of suicide attempt or death (n=302 missing such data were excluded from analyses). Of the total sample, n=1,086 had African ancestry (of whom 988 [97.1%] parent-reported as Black race and 71 [6.6%] parent-reported as Hispanic); and n=4,128 had European ancestry (of whom 4,093 [99.2%] parent-reported as White race and 123 [3%] parent-reported as Hispanic). The ABCD Study® protocol was approved by the University of California, San Diego Institutional Review Board (IRB), and was exempted from a full review by University of Pennsylvania IRB.

### Variables Exposures

#### Polygenic risk score of suicide attempt

ABCD Study genotype data was processed as described before (9). Polygenic risk scores were calculated separately for African- and European-ancestry participants using PRSice-2 (10), with summary statistics from a recent genome-wide association study (GWAS) of suicide attempters that included diverse ancestries (11). We tested eight P-value thresholds (Pt): 0.0001, 0.001, 0.01, 0.05, 0.1, 0.3, 0.5, and 1. Ten within-ancestry genetic principal components were regressed out of z-scored polygenic scores.

#### Parental history of suicide attempt

Parental history was evaluated using parent reports on parents’ suicide attempt/death (variable: “famhx_ss_momdad_scd_p”).

### Outcome measure

Self-report of suicide attempt in any of the three first ABCD Study assessments. Participants were considered controls if they denied history of suicide attempt in all three assessments.

### Statistical Analyses

Analyses were conducted from January-March 2022 using ABCD Study data release 4.0. Data preprocessing and analysis are detailed at https://github.com/barzilab1/ABCD_SA_genetics_FH.

Mean (standard deviation [SD]) and frequency (%) were reported for descriptive purposes. Univariate comparisons were made using t-test or chi-square tests, as appropriate. We used two-tailed tests for all statistical models. We imputed age for participants who did not complete the 2-year follow-up assessment (n=21, 0.4%). We used R version 4.1.0. for data analyses.

We estimated binary logistic regression models with suicide attempt as the dependent variable and PRS-SA as the independent variable, co-varying for age, sex and parent-reported race. A GWAS P-value threshold of P=0.05 was selected for the PRS-SA, to derive the highest Nagelkerke’s R^2^ and lowest P-value of PRS-SA in association with suicide attempt. We used a permutation test to validate this selection (see figure in the online supplement). Association of PRS-SA with suicide attempt was consistent across multiple P-value threshold tested (see table 1 in the online supplement).

**Table 1.** Sample characteristics.

|  | Total sample<br>N=5 214 | Control<br>n=5 032 | Suicide attempt<br>n=182 | P-value |
| --- | --- | --- | --- | --- |
| Age, years, mean (SD) | 12.04 (0.65) | 12.04 (0.65) | 12.09 (0.66) | 0.325 |
| Female sex, No. (%) | 2 408 (46.2) | 2 319 (46.1) | 89 (48.9) | 0.501 |
| Race Black, No. (%) | 1 087 (20.8) | 1 021 (20.3) | 66 (36.3) | <0.001 |
| Parent suicide attempt/death, No. (%) | 306 (5.9) | 279 (5.5) | 27 (14.8) | <0.001 |
| Suicide attempt PRS<sup>a</sup>, mean (SD) | 0.00 (0.95) | -0.01 (0.95) | 0.21 (0.89) | 0.001 |

<sup>a</sup> PRS after standardizing the raw PRS produced at a GWAS P-value threshold of 0.05 and then regressing out the first ten within-ancestry genetic principal components. Abbreviations: PRS= polygenic risk score; GWAS= Genome wide association study.

To test the additive effects of PRS-SA in explaining variance in suicide attempt, we estimated regression models with and without PRS-SA and compared the goodness of fit using the likelihood ratio test.

In sensitivity analyses, to address the potential bias of PRS-SA performance in African versus European ancestry, we estimated models stratified by ancestry and then meta-analyzed the results.

## Results

Among the 5,214 participants, 182 (3.5%) endorsed having made a suicide attempt at least once in the three ABCD Study assessments. History of suicide attempt was more frequent among Black youth (66 of 1,087, 6.1%) than among their White counterparts (116 of 4,127, 2.8%, Chi-square P<.001). No age or sex associations were observed. Participants who endorsed suicide attempt had more parental history of suicide attempt/death (14.8% versus 5.5%, respectively, Chi-square P<.001). **Table 1** includes univariate comparisons between participants with and without history of a suicide attempt.

PRS-SA was significantly associated with suicide attempt in the full sample (odds ratio [OR]= 1.3, 95% confidence interval [CI] 1.1-1.5, P=.001, co-varying for demographics: age, sex, parent-reported race, and accounting for ten within-ancestry genetic principal components). This model explained 2.7% of the variance (Nagelkerke’s R^2^=0.027), significantly more than the base model that only included covariates (Nagelkerke’s R^2^=0.019, likelihood ratio Chi-square test, P=.001). **Figure 1** illustrates the association between PRS-SA and suicide attempt rate.

**Figure 1.**
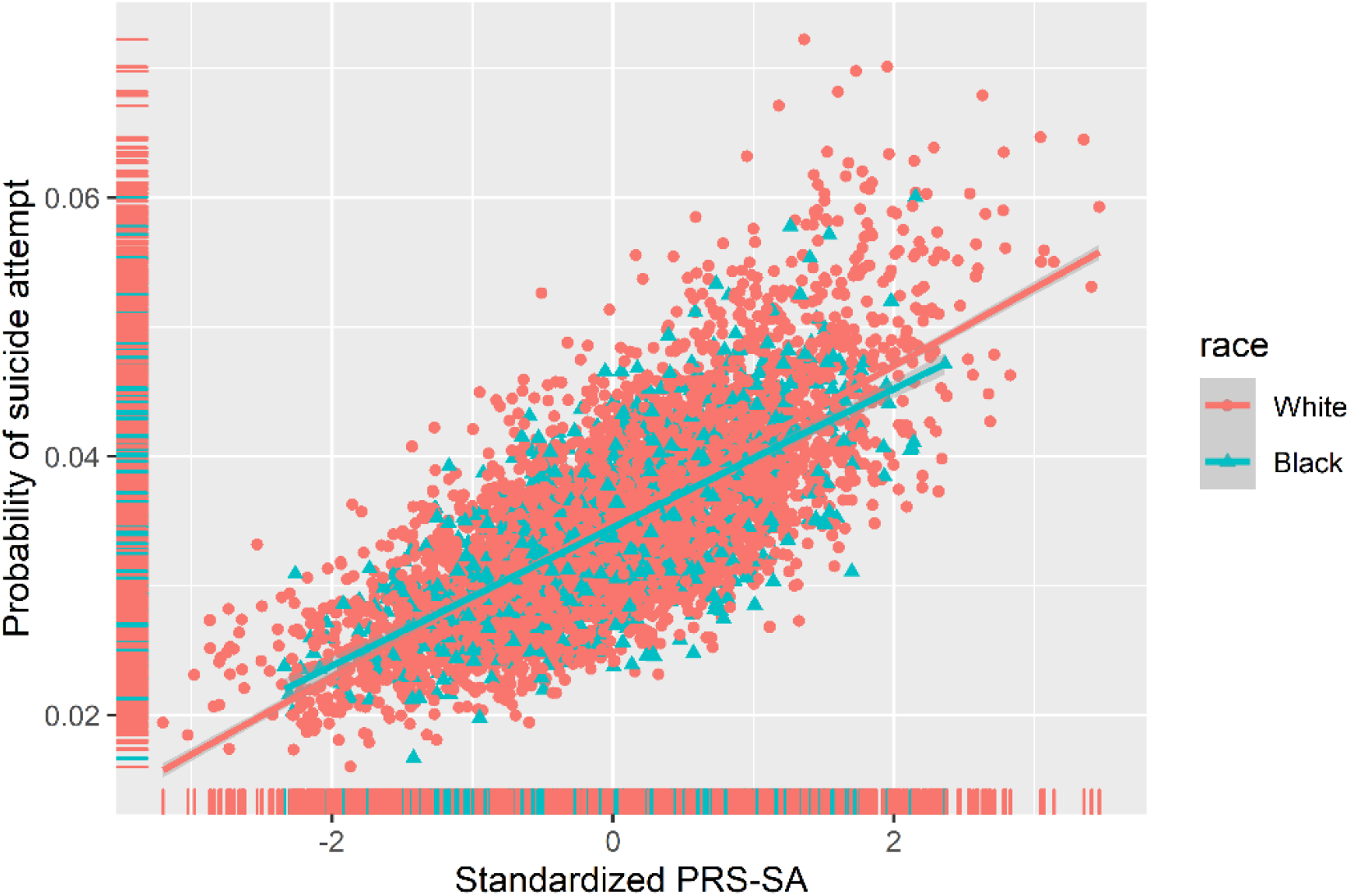
Polygenic risk score for suicide attempt (PRS-SA) and suicide attempt in Black and White youth. Scatter plots and regression lines show estimated probabilities of suicide attempt in 5 214 adolescents obtained from a binary logistic regression model with PRS-SA, age and sex as independent variables. X-axis represents PRS-SA score (after standardizing the raw PRS produced at a GWAS P-value threshold of 0.05 and then regressing out the first ten within-ancestry genetic ancestry principal components). Y-axis represents predicted probability of suicide attempt.

Parental history of suicide attempt/death was associated with youth’s history of suicide attempt (OR=2.9, 95%CI= 1.9-4.4, P<.001). Adding PRS-SA to this model increased the variance explained from 3.3% to 4% (likelihood ratio chi-square test, P=.002). The improvement in model performance (ΔR^2^=0.7%) obtained when adding SA-PRS was on the order of 50% of the ΔR^2^ obtained from adding parental history to the base model (ΔR^2^=1.4%, from 1.9% to 3.3%). Statistics for all models are detailed in **Table 2**.

**Table 2.**
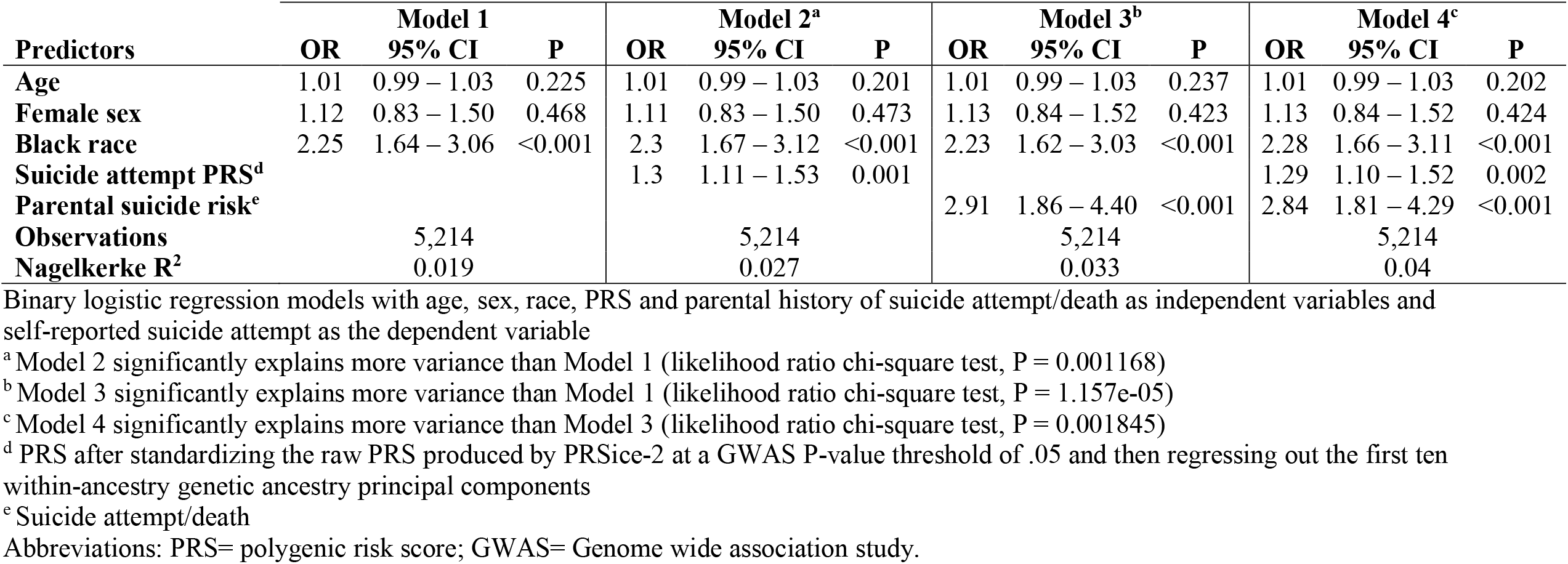
Association of suicide attempt PRS, parental history of suicide attempt/death and suicide attempt.

Finally, sensitivity analyses estimating the above models stratified by ancestry (African or European) and meta-analyzed yielded similar results in direction and statistical significance (see table 2 in the online supplement).

## Discussion

We present evidence suggesting clinical utility of a polygenic score explaining suicide attempt in Black and White US youth. Two main strengths of this work are noteworthy. First, the focus on suicide attempt highlights the clinical significance of the findings. Notably, most research in this age range lumps ideation and attempt together (12–14), even though most ideators do not make an attempt (3,15). Second, the inclusion of Black youth in the current work is critical to address racial disparities in psychiatric genetics research (16). This disparity is especially concerning in the field of youth suicide, where Black US youth are particularly vulnerable (2,17). Our findings extend recent ABCD Study results showing associations of depression polygenic risk score with suicide attempt in an analysis limited to European ancestry (18) and schizophrenia polygenic risk score with suicide attempt reported in the baseline ABCD Study assessment in admixed population with substantially fewer suicide attempt participants (64 versus 182 in the current analysis) (19).

We found that PRS-SA additively explains variance in suicide attempt beyond parental history of suicide attempt/death. *From a clinical perspective*, assessment of family history is common practice for clinicians to help their risk classification. We believe that clinicians can intuitively appreciate the value of PRS-SA when it is compared to this benchmark of clinical good practice. *From a research perspective*, considering skepticism in the field toward incorporating PRS in multivariable predictive algorithms in psychiatry (5), our findings provide support for incorporation of genetic scores, including that of suicide attempt, in suicide risk prediction (20).

One key limitation should be noted. The variance explained by addition of PRS-SA to models of parental history is still relatively small. We believe that with growing power/ sample sizes of GWAS in more diverse samples, the added value of PRS will increase, especially in diverse populations.

## Conclusions

In this cohort of young adolescents, PRS-SA was associated with suicide attempts and significantly improved models explaining variance over and above parental history of suicide attempt/death, which is commonly used in clinical settings to assess suicide risk. Findings suggest that PRS-SA may be useful for suicide risk classification in both Black and White youth.

## Data Availability

Data used in the preparation of this article were obtained from the ABCD Study (https://abcdstudy.org), held in the NIMH Data Archive.

https://nda.nih.gov/abcd/

## Acknowledgement

Data used in the preparation of this article were obtained from the Adolescent Brain Cognitive DevelopmentSM (ABCD) Study (https://abcdstudy.org), held in the NIMH Data Archive (NDA). This is a multisite, longitudinal study designed to recruit more than 10,000 children age 9-10 and follow them over 10 years into early adulthood. The ABCD Study® is supported by the National Institutes of Health and additional federal partners under award numbers U01DA041048, U01DA050989, U01DA051016, U01DA041022, U01DA051018, U01DA051037, U01DA050987, U01DA041174, U01DA041106, U01DA041117, U01DA041028, U01DA041134, U01DA050988, U01DA051039, U01DA041156, U01DA041025, U01DA041120, U01DA051038, U01DA041148, U01DA041093, U01DA041089, U24DA041123, U24DA041147. A full list of supporters is available at https://abcdstudy.org/federal-partners.html. A listing of participating sites and a complete listing of the study investigators can be found at https://abcdstudy.org/consortium_members/. ABCD consortium investigators designed and implemented the study and/or provided data but did not necessarily participate in analysis or writing of this report. This manuscript reflects the views of the authors and may not reflect the opinions or views of the NIH or ABCD consortium investigators.

## Author Contributions Statement

RB conceptualized the study question and study design, interpreted the findings and wrote the first draft of the manuscript. EV curated and processed the phenotypic data and conducted data analysis. LS processed all genomic data, calculated polygenic risk score and supervised statistical analyses. VW and ND substantially helped in study conceptualization, data interpretation and preparation of the first draft of the manuscript. LA supervised study conceptualization and all statistical analyses and contributed to data interpretation. All authors made substantial contribution to editing and revising the manuscript to its final version.

